# Self-reported adaptability among postgraduate dental learners and their instructors: accelerated change induced by COVID-19

**DOI:** 10.1101/2021.06.05.21258401

**Authors:** Farah Otaki, Fatemeh Amir-Rad, Manal Al-Halabi, Zaid Baqain, Nabil Zary

## Abstract

It is forecasted that the skills and competencies necessary for post-pandemic success in higher education need to be founded upon adaptability, coping, and Self-regulated Learning (SRL). It is worth investigating how stakeholders perceived their adaptability and coping with the accelerated change accompanying COVID-19. Accordingly, the purpose of this study was to assess the self-reported adaptability of postgraduate dental learners and their instructors in the context of abrupt transition to distance learning induced by the pandemic.

This study utilized a convergent mixed methods study design. The qualitative and quantitative data were concurrently collected from instructors and learners. The datasets were analyzed independently, and the generated information was integrated using a joint model analysis.

The percentage of average of self-reported adaptability of both groups was 81.15%. The instructors, with a mean of satisfaction of 17.94 (±1.76), rated their adaptability significantly higher than the learners, with a mean of satisfaction of 15.66 (±2.77) (p=0.002). The thematic analysis resulted in two interrelated themes: Self and Environment. Within the Self theme, three subthemes surfaced: Cognitions, Emotions, Behaviors. As for the Environment theme, it encapsulated two subthemes: Enablers and Impediments.

The stakeholders perceived themselves to have adapted well to the transition, and SRL appeared as a cornerstone in the adaptation to the accelerated change (accompanying COVID-19). There appeared to be an interplay between the cognitions, emotions, and behaviors on the level of the self as part of the adaptation process. Also, building upon existent models of SRL, this study uncovered that the stakeholders considered the environment to play a crucial role in their adaptation process. This highlights the importance of developing a climate that remains, despite external pressures, conducive to attaining learning and teaching goals. It is also crucial for university-level mental health promotion activities to proactively foster, among learners and instructors, adaptability, building ‘academic resilience’.

## Introduction

The COVID-19 pandemic made characterizing today’s world as Volatile, Uncertain, Complex and Ambiguous (VUCA) more relevant than ever before (1-3). This pandemic brought about an accelerated change where remote interaction became the only plausible solution at a point in time. The need to “go remote” at the onset of the pandemic accelerated innovation in telecommunication. It brought to the forefront the previously underused internet-based services and products, such as telehealth (4), e-commerce (5), and distance learning (6).

This accelerated change was evident in higher education (6), where learning and teaching all around the world had to switch to the online environment abruptly (7). The volatility of the environment got heightened due to the continuous changes that this sector is having to keep up with (8). This is associated with uncertainty. Although a lot of research and investigations are taking place to enable foresight (9), no one knows with any great certainty the current and long-term effect of the pandemic on learning and teaching. The situation has been novel and seemingly uncontrollable and remains unresolved (10). The introduced complexity has been evident on all socioecological levels of higher education, where stakeholders need to deal with diverse stressors (including but not limited to: safety concerns, sense of isolation and loneliness, and complete disruption of daily routines), along with mental health difficulties such as isolation and loneliness (10), and depression and anxiety (11). Since these times are unprecedented, there is a substantial amount of ambiguity that all the higher education stakeholders need to deal with (11); everyone appears to be resorting to trial-and-error techniques to adapt (12).

This situation revealed how inadequately prepared higher education was for radical transformation (13-16). This is especially true in health professions education, where educators got challenged with ensuring the protection of the health and wellbeing of all their stakeholders, including the trainees, continuity of quality education, confidence in health and safety measures, and abidance to guidelines for clinical training. Many teaching clinics and academic hospitals ended up suspending all activities involving trainees. This significantly affected students’ quality of education, where their experiential learning and clinical exposure got compromised considerably (17). If not made up for, this gap in clinical learning will inevitably impact developing the competencies required for nurturing and graduating safe, competent healthcare providers (18).

It is forecasted that the skills and competencies necessary for post-pandemic success in education during the next five years need to be founded upon adopting skills related to emotional intelligence, learning and innovation, and information, media, and technology (19-21). Accordingly, from an educational psychology perspective, it is essential to highlight Self-regulated Learning (SRL) and its constituents. Systemically leveraging the cognitive, metacognitive, behavioral, motivational, and emotional/affective aspects of learning becomes more important as the external environment becomes more challenging. This, in turn, will raise their self-efficacy and sense of agency, cognitive resilience, and adaptability (22). In fact, from this perspective, adaptability, coping, and SRL becomes intertwined. Adaptability has been conceptualized and defined as “… the capacity to constructively regulate psycho- behavioral functions in response to new, changing, and/or uncertain circumstances, conditions, and situations…” (23).

It is worth investigating how well the relevant stakeholders perceived themselves to adapt and cope with the accelerated change, and the corresponding VUCA, accompanying COVID-19. Accordingly, the purpose of this study was to assess the self-reported adaptability of postgraduate dental learners and their instructors in the context of abrupt transition to distance learning induced by the onset of the pandemic. The study’s research questions were as follows:

- How adaptable did the stakeholders under investigation perceived themselves to be?
- How did the stakeholders adapt their approaches (be it in relation to the learning and teaching, or otherwise) to cope with the accelerated change?
- How can other postgraduate dental schools proactively raise the level of adaptability of individuals and the all-encapsulating institutions?

## Materials and methods

### Context of the study

This study took place at the Mohammed Bin Rashid University of Medicine and Health Sciences (MBRU), Dubai, UAE in the Hamdan Bin Mohammed College of Dental Medicine (HBMCDM). This postgraduate dental college offers three-year full-time specialty dental postgraduate programs in prosthodontics, periodontics, pediatric dentistry, orthodontics, and endodontics.

### Responding to COVID-19

HBMCDM, along with all other educational institutions in the UAE, switched to complete distance learning from 22^nd^ March 2020 until the end of the respective academic year. Despite all the surfacing impediments, HBMCDM stakeholders managed to continue all didactic educational activities as previously scheduled (6).

### Transitioning to distance learning

Two digital platforms were utilized to deliver distance learning, the Learning Management System (LMS) and Microsoft Teams. These platforms enabled real-time class presentations, research dissertation-related communications, and clinical case-based discussions (CBD). In addition, some instructors pre-recorded their lectures for learners to access and ‘consume’ the content at their convenience.

MBRU Faculty Development and Information Technology (IT) support teams conducted training sessions and designated technical support personnel to assist the faculty throughout the transition. Although a previously set schedule for the delivery of all didactic courses was maintained, the instructors limited the sessions’ length to one hour each. They provided reading material before the teaching sessions to deliver the lessons’ intended learning outcomes within the shortened duration. Class attendance was registered on MBRU Self-Service portal. To partially compensate for the lack of clinical learning due to the suspension of clinics, CBD was delivered on MS Teams across two four-hour sessions per week.

Due to the absence of live proctoring and the inability to conduct final examinations on-campus, instructors were encouraged to consider alternative assessment methods that test the attainment of the courses’ learning outcomes and hold the learners accountable to academic integrity. Instructors were encouraged to conduct assessments orally using clinical scenarios, especially in complex multidisciplinary cases via MS Teams. The LMS system, through which written exams were conducted, adopted a lockdown browser requirement preventing access to any other application during the exam. Also, activation of the webcam, in the learners’ devices, was required. Adequate training and support were provided to both learners and instructors on using the additional exam security requirements mentioned.

### Research design

The study’s ethical approval was granted by the MBRU, Institutional Review Board (Reference # MBRU-IRB-2020-032). As part of a multi-phased research project, this convergent mixed methods study design (24) was adopted to develop a thorough understanding of the extent of adaptability of the learners and their instructors during the rapid transition to distance learning due to COVID-19. To start with, the qualitative and quantitative data were concurrently collected (from both groups of stakeholders: instructors and learners). The quantitative datasets were analyzed independently from the qualitative datasets. Then, the generated information was integrated relying on joint model analysis (25, 26).

### Data collection

The data was collected using a survey that was designed specifically for this research project. The survey was composed of two segments. The first segment constitutes four components measured against a Likert-type scale of five points (1: Strongly Disagree, 2: Disagree, 3: Neutral, 4: Agree, and 5: Strongly Agree). The 4 components are as follows:

1. I was able to effectively cope with the higher technological demands of distance learning.
2. I was able to manage my time and efforts to cope with the transition to distance learning.
3. I was able to monitor and evaluate my performance, and if need be- intervene, to cope with the transition to distance learning.
4. I sought help, if needed, from students, colleagues, University staff, and/ or family members to cope with the transition to distance learning.

The second segment of the survey was meant to be exploratory to solicit for qualitative data using the following open-ended questions:

- How did you feel about the transition to distance learning at the beginning?
- How do you feel about the transition to distance learning now that some time has elapsed?
- How did you cope with the transition to distance learning (related to the learning experience, or otherwise: personal-level, environment, and friends and family)?
- Reflect on the changes to your teaching approach due to the transition to distance learning.

All full-time and part-time faculty involved in the distance learning at HBMCDM were invited to participate. Students from the five different postgraduate programs in all three levels were also invited to participate. No personal identifiers were recorded to ensure the privacy and confidentiality of all participants. The participation in this survey was completely voluntary. The survey was open for learners and instructors for participation from May 1^st^ through 31^st^ 2020. At the time of data collection, HBMCDM had 21 instructors and 63 students. The faculty was composed of 5 females and 16 males, with an average age of 48 years. The faculty were of 12 different nationalities, with the following distribution of academic rankings: 4 professors, 5 associate professors, 5 assistant professors, and 7 lecturers. Nineteen were full-time and 2 were part-time employees. The learners consisted of 44 females and 19 males, distributed across 16 different nationalities, with an average age of 30.9 years.

The Strategy and Institutional Excellence team at MBRU (i.e., the unit handling the Quality Assurance and Institutional Effectiveness portfolio) sent an email, with the survey link, to all participants on May 1^st^. Weekly email reminders were sent out until the closing of the survey on May 31^st^. Also, the Dean of HBMCDM sent emails to both the faculty and students’ groups independently to highlight the importance and value of voicing one’s opinion through participating in this survey.

### Data analyses

#### Quantitative analysis

The quantitative data was descriptively analyzed using SPSS for Windows version 27. For each of the four quantitative components, the mean and standard deviation were generated. An overall score of adaptability was calculated.

Since the scale used for capturing the perception of the learners and instructors was tailor-made for this study, the validity tests of Cronbach’s Alpha and the Principal Component Analysis (PCA) were performed to ensure internal consistency and check external variance, respectively, of the adapted tool.

To select the appropriate inferential analysis test, a test of normality was conducted for each of the four components and for the score of adaptability. The data of each of the four components, independently, and the score of adaptability all turned out to be not normally distributed. Accordingly, the Mann-Whitney test was used to compare the overall score of adaptability, and each component independently, between both groups of stakeholders (learners and instructors).

#### Qualitative analysis

The qualitative data analysis started after the completion of the data collection phase. The data was inductively analyzed using thematic analysis. The process of analysis followed Braun and Clarke (2006) (27) six-step framework, which is a recommended approach for thematic analysis in health professions’ education (28). The indicated six steps for conducting thematic analysis include: 1- familiarizing oneself with the data, 2- generating initial codes, 3- searching for themes, 4- reviewing themes, 5- defining and naming themes, and 6- producing the report.

NVivo software version 12 plus (QSR International Pty Ltd, Vic, Australia) was used to code the data, and in turn, facilitate the categorization of the relevant text fragments. The data collected from each of the two groups of stakeholders was handled separately.

First, two researchers (FO and FAR) familiarized themselves with the data by examining and re-examining the qualitative data. Second, the qualitative, narrative data was examined line-by-line and initial codes were generated. The two researchers underwent several rounds of discussions to resolve any discordances between their observations. Third, the initial codes were inductively investigated to be combined into subthemes, which in turn went through a similar iterative process to construct themes. The generated themes and sub-themes were then reviewed as part of stage four to ensure that the data within each compartment (i.e., theme or subtheme) are sufficiently common and coherent, also the compartments are adequately distinct from each other. As part of stage five, themes and subthemes were coded and defined in the context of the study. The last step constituted of reporting upon the findings of the qualitative analysis.

#### Mixed Methods Integration

After completing the independent quantitative and qualitative analyses, the outputs were systematically integrated via joint display analysis. This involved merging the results from the two datasets through a side-by-side comparison to assess the best way to map the findings onto each other. This iterative process enabled developing a whole that is greater than the sum of its parts (24, 29). As such, the convergence of findings led to the development of a better understanding of the adaptability of learners and instructors during the transitioning to distance learning.

The Pillar Integration Process (PIP) framework, initially presented by Johnson et al. (2017) (30), was selected as the foundation of the adapted joint display analysis. The four stages of PIP were completed sequentially: 1- presenting the quantitative raw data and findings on the left side of the display, 2- exhibiting the qualitative raw data and findings on the right side of the display (establishing links between both sides of the display where possible), 3- confirming that both sides of the display match each other (as much as possible), and 4- “pillar-building” which is the ultimate stage of this process leading to the generation of meta-inferences which are position in the center of the display. As such, areas of data confirmation (where findings from both datasets reinforce each other) and data expansion (where a finding from one type of analysis is unique and has no match in the other type of analysis, but rather expand upon it by establishing complementarity) were identified.

## Results

The final respondents’ number was 53 out of 63 learners (i.e., response rate was 84%). As for the instructors, 18 faculty members out of 21 responded (i.e., response rate was 86%). A unique identification number was given to each of the 71 participants. It was complimented with ‘R’ for the learners and ‘I’ for the instructors (i.e., participants 1 through 53 were followed by ‘R’, and 54 through 71 by ‘I’).

### Quantitative analyses

The reliability score of Cronbach’s Alpha for the evaluation instrument, which captured the stakeholders’ perception, was 82.4%. The percentage of the total average of the learners, instructors, and both groups of stakeholders were 78.3%, 89.7%, and 81.15%, respectively, as per Table 1.

**Table 1.**
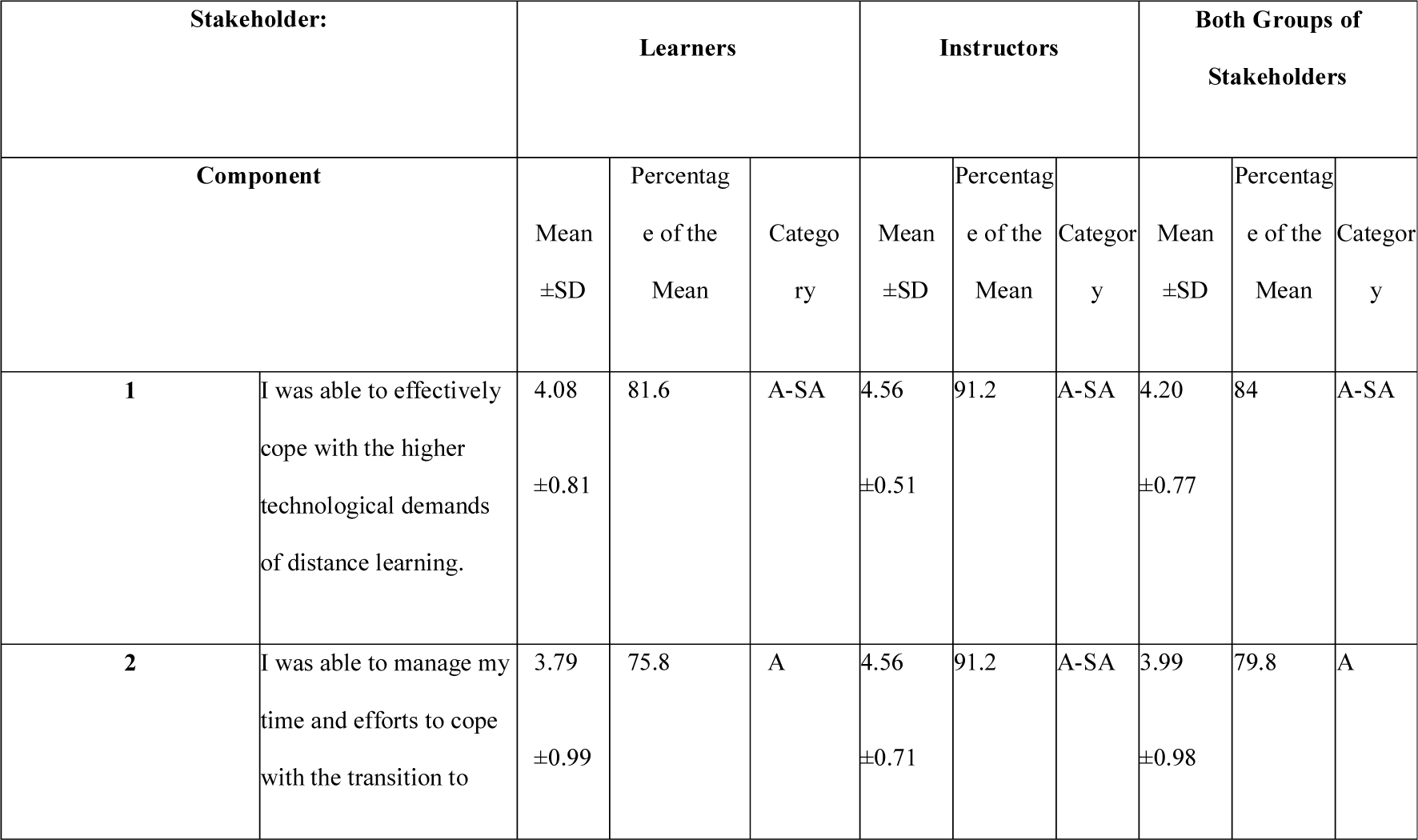
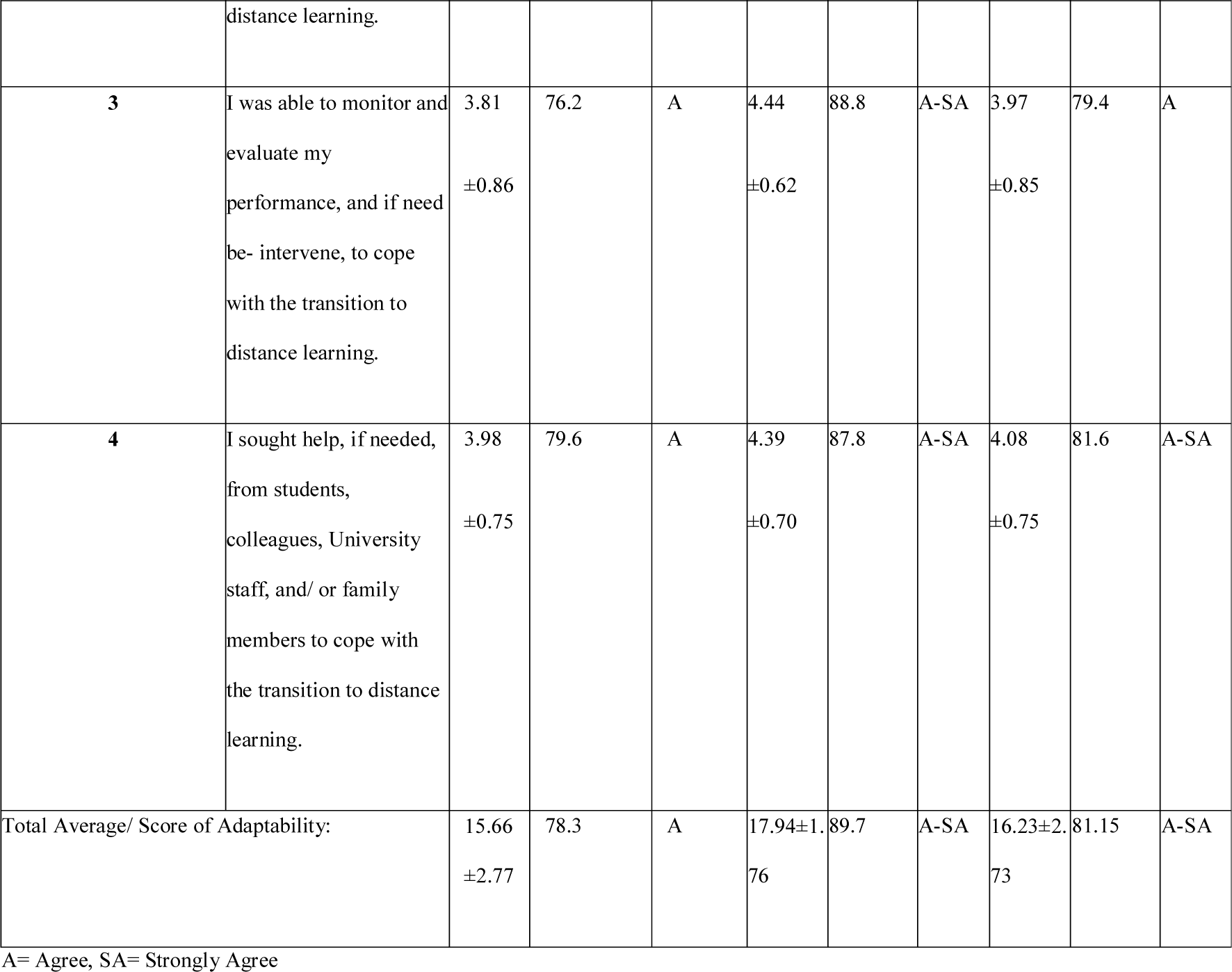
Output of descriptive quantitative analysis

According to the PCA, 87.1% of the variance can be explained by the instrument, which means the instrument is reliable and valid to measure what it is intended to measure.

As illustrated in Fig 1, the instructors, with a mean of satisfaction of 17.94(±1.76), rated their adaptability higher than the learners, with a mean of satisfaction of 15.66(±2.77) (p=0.002).

**Fig 1.**
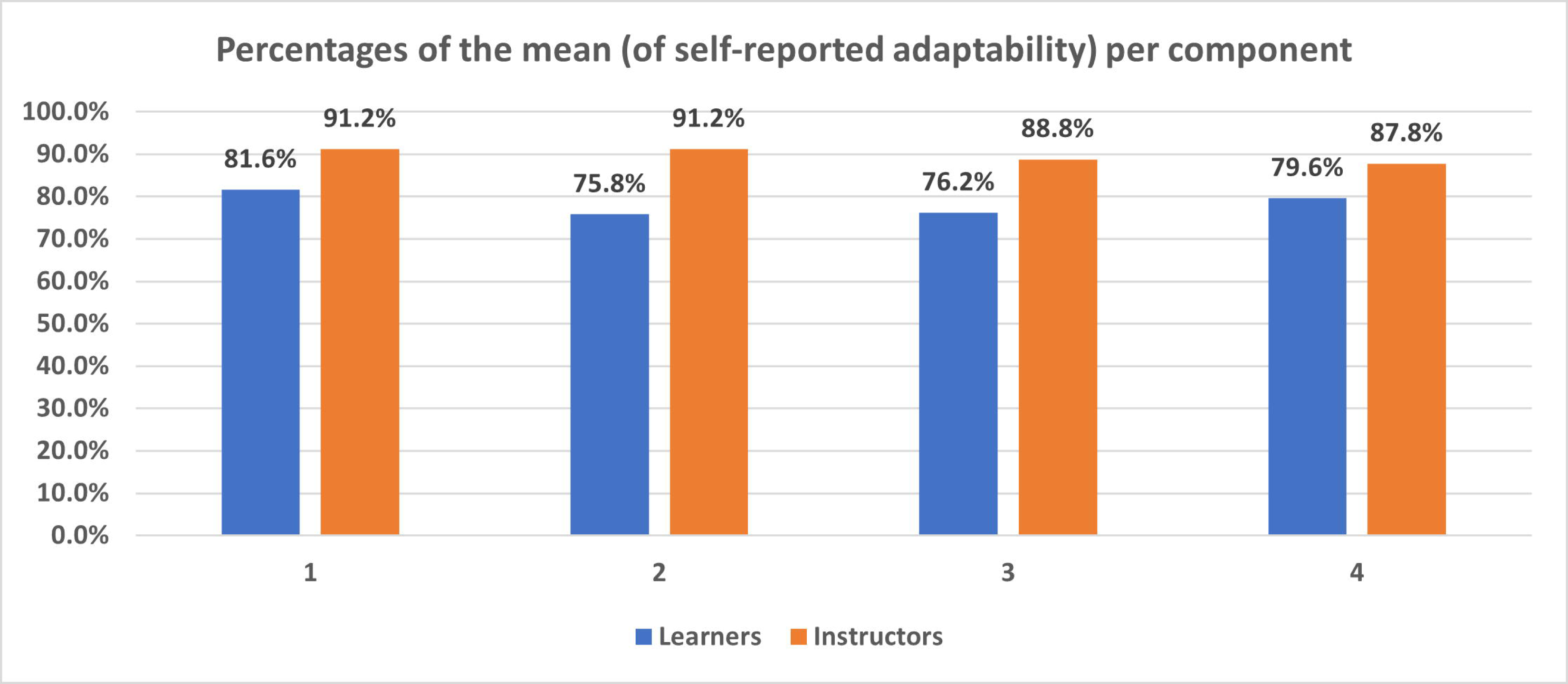
Comparison between percentages of the mean per component (1 through 4) between learners and instructors

### Qualitative Data

The thematic analysis resulted in two interrelated themes: Self and Environment, as illustrated in this study’s conceptual framework (Fig 2). Within the Self theme, three interrelated subthemes surfaced: Cognitions, Emotions, Behaviors. As for the Environment theme, it encapsulated two subthemes: Enablers and Impediments.

**Fig 2.**
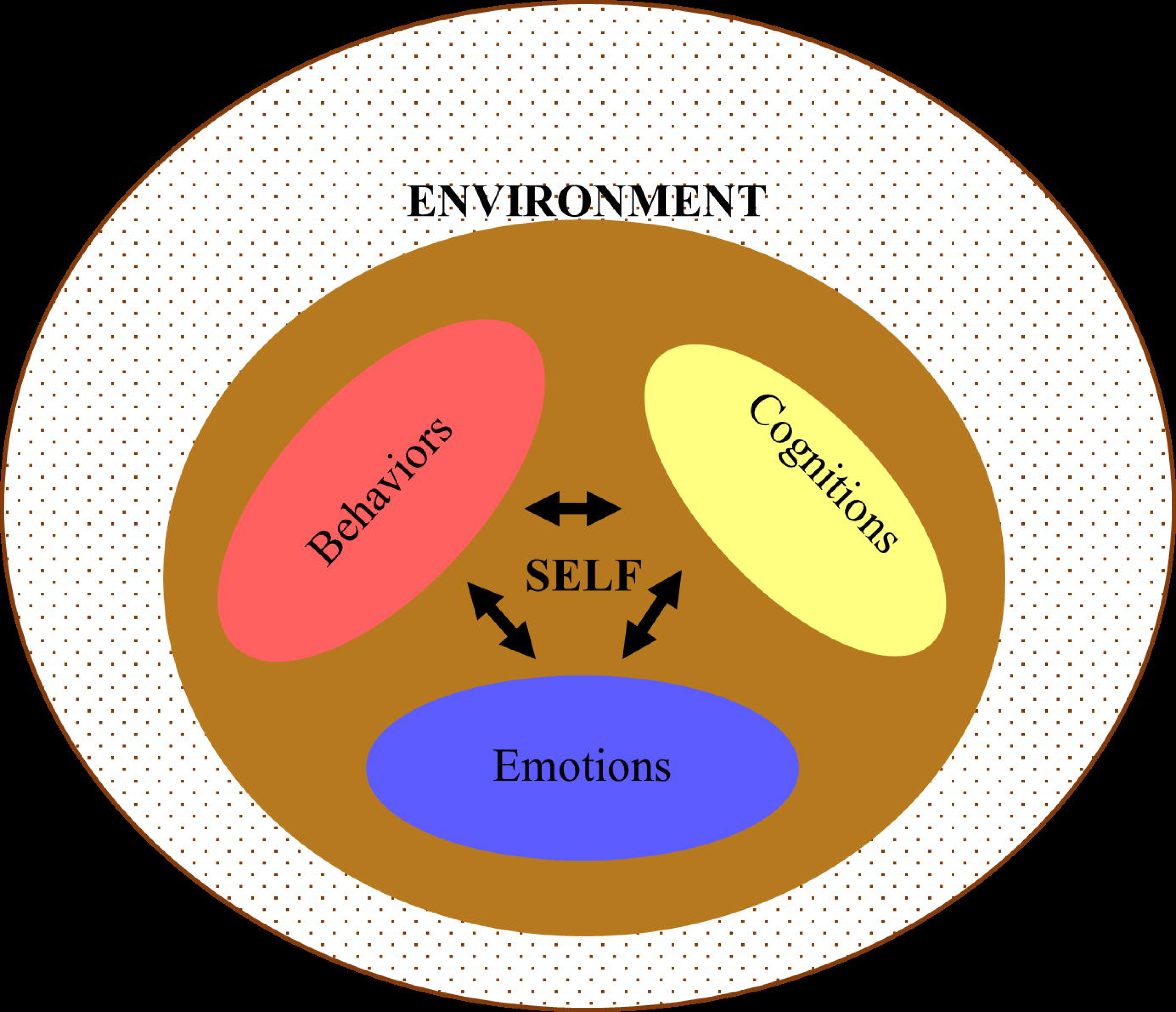
Study’s Conceptual Framework

#### Theme 1: Self

This theme focuses on intrapersonal factors that appeared to influence the learners’ and/ or the instructors’ adaptability.

##### Cognitions

This subtheme refers to thought processes that the individual learners and instructors seemed to be experiencing. These thoughts appeared to be related to oneself, other people, and/ or the context of experience.

###### Oneself (including meta-cognitions)

The stakeholders appeared to be judging themselves. They have somehow developed an opinion about how well they adapted to the situation.

L-9: “…I adapted and performed much better than I expected…”

I-19: “…I believed in myself; I was confident about my abilities to successfully transition to distance learning….”

In many instances, the learners and instructors appeared to be interpreting their own thoughts and understandings during the transition to distance learning.

L-10: “…I could not stay focused for long periods of time…”

L-31: “…most of the time, I was not able to focus my thoughts. I frequently experienced brain fogs….”

I-11: “…I was quite confused, at the beginning. For example, I was not sure if I needed to look at the laptop camera or at the screen where my presentation was shared from my desktop…”

###### Other people

The study participants reflected upon their own thoughts and opinions about others.

L-5: “…not sure if other students were paying attention during the lectures…”

I-11: “…at the beginning, I was doubting the students’ engagement…”

I-12: “…I think we became more interactive, over time. At the beginning, it was a challenge to both groups of stakeholders: the instructors and students. Eventually, we comprehended what a virtual classroom is…”

###### Context of experience

The participants also discussed their views around aspects related to their environment and the context of the experience. Some learners perceived the distance learning experiences to be difficult:

L-16: “…I found the experience to be quite challenging; I could not concentrate at home. It is not the right place to attend a class from…”

L-46: “… distance learning is sort of different from that which happens on campus in terms of motivation and interactions…”

I-11: “…the idea of finding myself on my own in my room, interacting with a screen was quite challenging…”

Others showed openness to and acceptance of the new experiences. They were grateful to the fact that the virtual environment enabled the continuation of learning in a time when in- person activities became unsafe.

L-9: “…it has been a pleasant new experience… I was intentional about adapting to the reality of the situation. I perceived it to be to our own benefit…”

L-45: “…at the beginning I was wondering how it will be. Later, I got surprised by how smooth the transition was…”

I-7: “…the distance learning was the best available alternative…”

I-9: “…the online teaching experience bought with it plenty of new opportunities. It has broadened the scope of learning and teaching…”

#### Emotions

This theme sheds light on the emotions that the participating learners and instructors experienced in adjusting to the distance learning and teaching. Some emotions were positive, and others were negative.

##### Positive (including motivational affects)

The study participants expressed several positive emotions:

L-7: “…I felt excited about trying this new experience…”

I-19: “…I am happy about all facets of distance learning…”

##### Negative

The stakeholders also expressed negative emotions. Its sudden onset and all the uncertainty around it were quite disturbing to some learners and instructors.

L-17: “…I was doubtful…”

I-6: “…I felt slight apprehension…”

I-8: “…I felt skeptical about the feasibility of the transition and maintenance of distance learning. How are we supposed to acquire the needed skills virtually? We are meant to become clinicians after all…”

Several learners referred to an elevation in their level of anxiety. Besides worrying, the learners also expressed anger, frustration, and changes in energy levels. Between caretaking, housework, kids’ homeschooling, and learning, some found personal and professional time blended.

L-2: “…I experienced feelings of frustration and exhaustion- there were many competing responsibilities, all taking place at home…”

L-3: “…I felt confused. Also, I got frustrated due to the many internet-related problems that I faced. This led to time management issues…”

L-46: “…I felt anxious, but I had trust that MBRU will find a way around the challenge, and will continue on providing us with the best…”

### Behaviors

This theme encapsulates all the different actions that the stakeholders partook to adapt to the change. It also included all the skills needed to exercise to keep up with the challenges inherent in the transition. The stakeholders differentiated between the behavioral changes that turned out to be conducive and those that appeared to them not to add any value to their experiences.

#### Constructive

Among the actions that were deployed, some turned out to be to the advantage of the learners and instructors. Some learners proactively developed opportunities to connect with peers; their engagement in virtual study groups added value to their learning strategies during the transition to distance learning.

L-14: “…I developed the habit to regularly meet with my colleagues online to study the lectures together…”

L-15: “…participating in group study was one of the best ways to keep distance learning as similar as possible to that of typical conditions….”

The learners also developed and deployed their time management skills.

I-7: “…I had to adjust my schedule to make better use of my time…”

Also, some learners were intentionally more engaged in the virtual context, relative to face- to-face interactions.

L-47: “…I started reading and researching more. I also developed the habit to prepare for the lectures, before the actual time the classes take place…”

L-7: “…I focused on preparing for the lecture ahead of time, and I maximized my participation during the meeting. It was clear that our instructors were trying their best to make-up for the interpersonal gap. When I have a lecture to present, I try to have to have pauses every now and then with a funny slide or so to refresh the energy of my colleagues…”

Instructors adjusted their teaching strategies to engage learners. They adopted techniques to foster meaningfulness in their connection with the learners and overcome physical and emotional isolation.

I-11: “…I started to share more links, videos, and reading materials with my residents to further support their learning. We arranged for online meetings on Teams to discuss their research projects, and address and reflect upon inquiries related to their presentation…”

I-12: “…our residents stayed in touch via WhatsApp groups that were created during the lockdown. It made connecting with and updating each other easier…”

Some stakeholders realized that they were proactive in modifying their physical environment at home for it to become more conducive to their learning and teaching targets.

L-22: “…I arrange a studying set-up at home… I made the effort to change this set-up from time to time…”

I-11: “…allocating a working space at home was also very helpful. I get into my work mode as soon as I land on this desk…”

#### Futile

In their attempt to cope with the transition, some stakeholders resorted to behaviors that did not add value to their experience. Learners were suddenly faced with many responsibilities that they needed to attend to concurrently from the same space. A few of those learners seemed to deal with all their responsibilities as one chunk, without any sort of compartmentalization.

L-12: “…It has been difficult to suddenly be required to manage both family and university at once, in the same place…”

Other learners could not bear the fear and uncertainty inherent in the situation and ended up overworking themselves as a coping mechanism.

L-7: “…the transition and isolation, and all the accompanying stress and fear are making me spend most of the time studying, which is getting quite stressful. I barely have any time left for myself. What I used to finish in one hour, now takes me 3 hours…”

Some learners needed to lessen their level of interactions to deal with their anxiety and insecurities.

L-24: “…I was hesitant to participate in the class unlike in the normal class setting I would have been more interactive…”

### Theme 2: ENVIRONMENT

This theme refers to external factors that the stakeholders perceived to have affected the learning and teaching experience positively or negatively.

#### Enablers

Among those external variables that the stakeholders shed light on were ones that inspired, enabled, and/ or empowered the learners and their instructors. For example, the stakeholders particularly emphasized that the understanding of family members was crucial for effectively transition.

L-21: “…the most important thing that my family understand the situation and do not interrupt me during my classes…”

Also, relying on family and friends for support and comfort was frequently brought-up by the study participants. The role of close family members appeared to be quite helpful in facilitating the individual-level adjustments that needed to take place.

L-47: “…it was not very difficult to cope on my own; my family was supporting and comforting me all the time…”

I-9: “…I coped well; thanks to good friends and family…”

In some cases, the instructors shed light on how the fact that they had rapport with the students enabled and smoothened the transition.

I-11: “…I think it is more convenient to move to distance learning after knowing the residents and they get to know you through face-to-face interaction…”

The participating stakeholders highlighted that one of the significant external resources was the educational institution itself. This included how the institution led the transition and provided all different kinds of support at various institutional levels to both learners and instructors.

L-24: “…the university was very cooperative…the instructors made me feel at ease, my colleagues kept on sharing with me stories of the obstacles that they had to go through and how they overcame them….”

I-12: “…the transition has been managed professionally by MBRU leaders and staff members, along with faculty members and other stakeholders who are directly involved in the learning and teaching. I perceive the transition to have happened smoothly…”

I-15: “…transition was a lot easier than I expected; thanks to Information Technology team support…”

#### Impediments

The stakeholders also identified external variables that they felt held them back, discouraged them, and/ or slowed them down. For example, learners indicated the challenge of the competing responsibilities that arose because of the pandemic and that needed to be attended to concurrently from the same physical space. They needed to strike a balance between keeping up with their educational duties and their personal and/ or familial life.

L-2: “… personally, I have no free time to read any supplementary material assigned by my instructors. I am always busy; we have to be available 24 hours a day, 7 days a week for anything or everything at home…”

L-3: “…it was quite challenging in the beginning. I could not continue having a part- time homecare nurse to support me with taking care of my father. Her part-time constituted a risk in terms of transferring the virus in between her multiple work duties…”

L-38: “…All of a sudden, I needed to keep an eye on my kids of 5 and 3 years of age, along with homeschooling them, while living-up to my learning obligations…”

L-25: “…I disliked it! I am a mother and having my kids around does not make it easy to focus. My kids need home schooling and supervision while I am having my classes…”

Some stakeholders considered that the lack of opportunities of hands-on and clinical experiences constitute a hindrance or obstruction towards their learning or teaching objectives.

I-2: “…in some areas it was ok, but others require interaction with students and hands-on experience…”

L-5: “…I miss my clinical work and patients, which is demotivating me…”

The sudden transition to the online environment accompanied by the isolation due to the social distancing directives constituted to almost all participants an external challenge that they needed to overcome. Some of the learners mentioned the loss of connection with others due to isolation. There were also the inevitable adverse effects of using electronic devices for long periods, which caused digital eye strain and/ or headaches among the learners.

L-14: “…it was hard to study at home sometimes, due to isolation. Studying on my own without my colleagues really affected me…”

L-14: “…I noticed that I was regularly experiencing headaches while studying from home, which was not the case prior COVID-19. These episodes were maybe induced by the heavy reliance on technologies and electronics…”

Disruption of daily routine and its consequences were repeatedly referred to by the learners and instructors. The switch to digital learning also resulted in sleep deprivation among our stakeholders. This, in turn, generated fatigue and in some cases burnout; the stakeholders observed that they were stretching themselves too thin.

L-2: “…the changes to my routine and increasing responsibilities were accompanied with lack of sleep. My lifestyle has not been healthy and there is very little that I can do about it…”

L-7: “…my personal space got conquered; I do not have the time now to recharge my own energy…”

I-5: “…not having a real break. It can get really tiring. With the lectures and meetings, I feel like I have been working 24/7…”

### Mixed Methods Integration

The PIP joint display visually conveys the inferences of the quantitative and qualitative analyses and meta-inferences generated by merging the outputs of both analyses. As depicted in Table 2, it is evident that the stakeholders perceive themselves to have adapted well to the transition, where the qualitative and the quantitative output of data analyses confirm each other. Relevantly, the quantitative analysis also revealed that the instructors perceive themselves to be adaptable significantly more than how adaptable the learners perceive themselves to be. These findings appear to complement the qualitative findings that there is an interplay between the cognitions, emotions, and behaviors on the level of the self as part of the adaptation process. Also, on its own, the qualitative findings shed light on the fact that the stakeholders considered the environment to play an essential role in their adaptation process, where they pinpointed enablers and impediments.

**Table 2.**
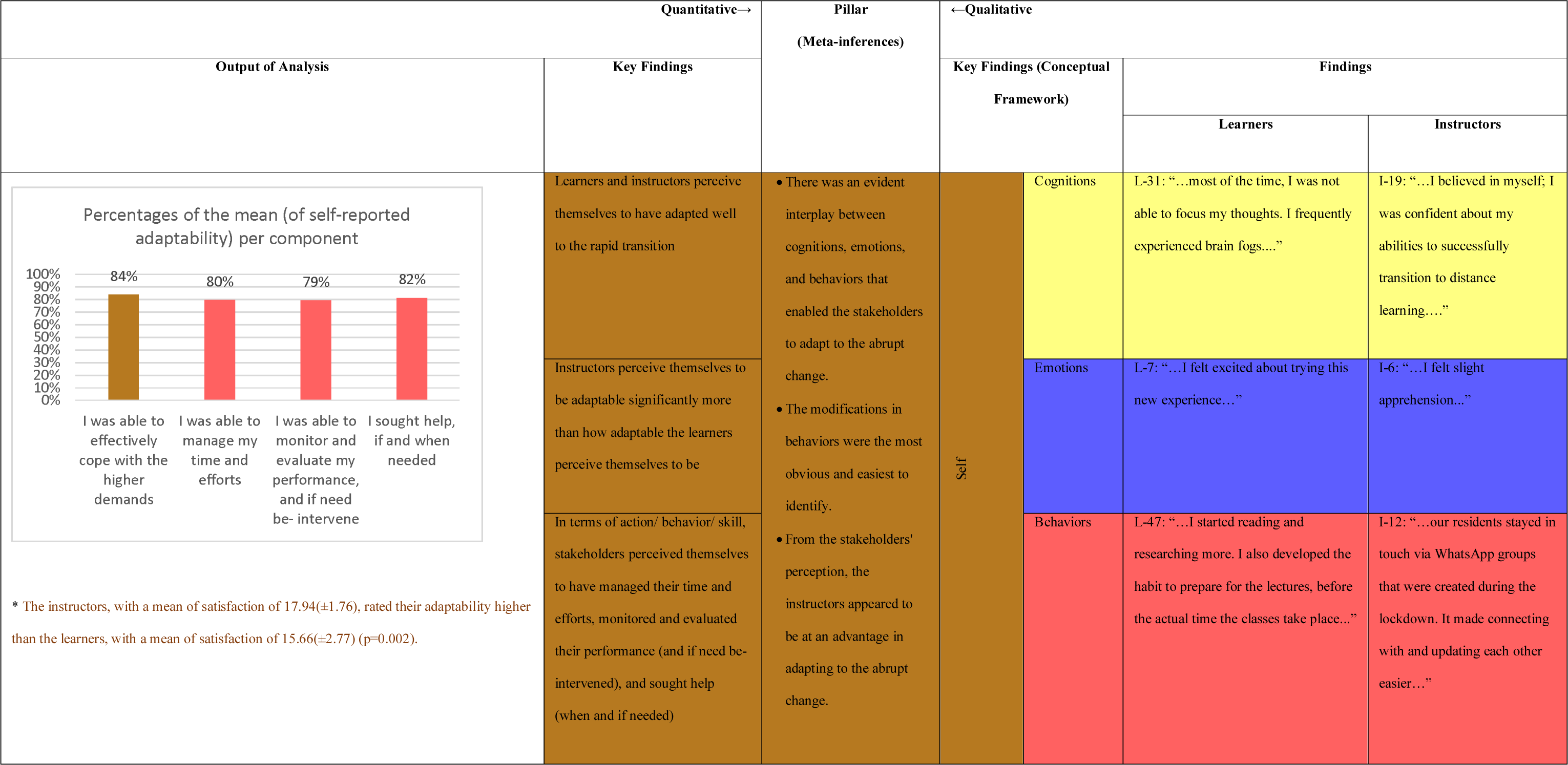
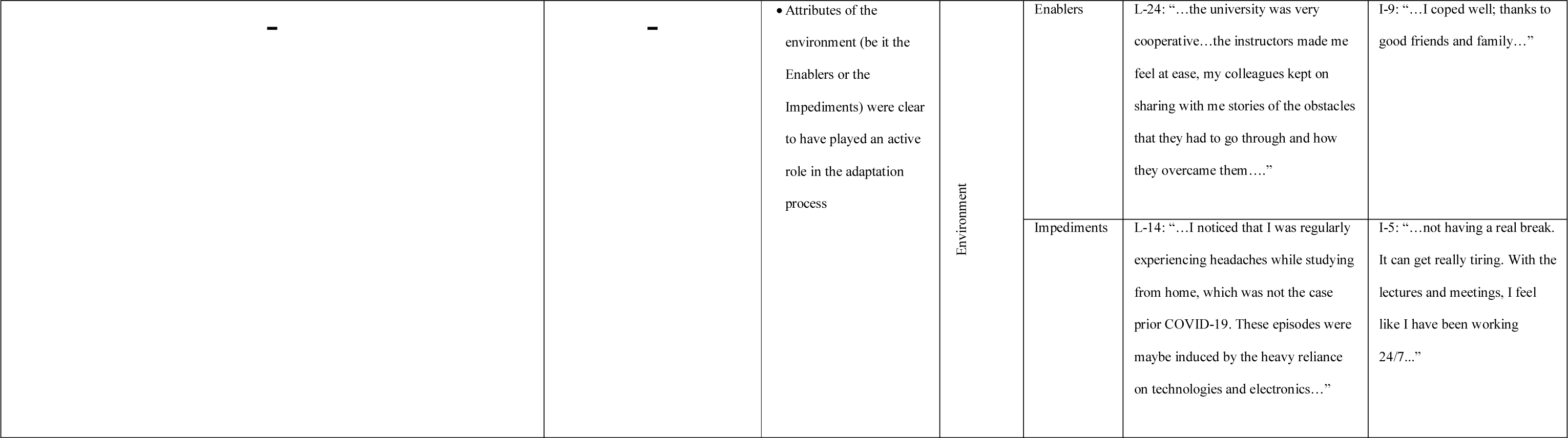
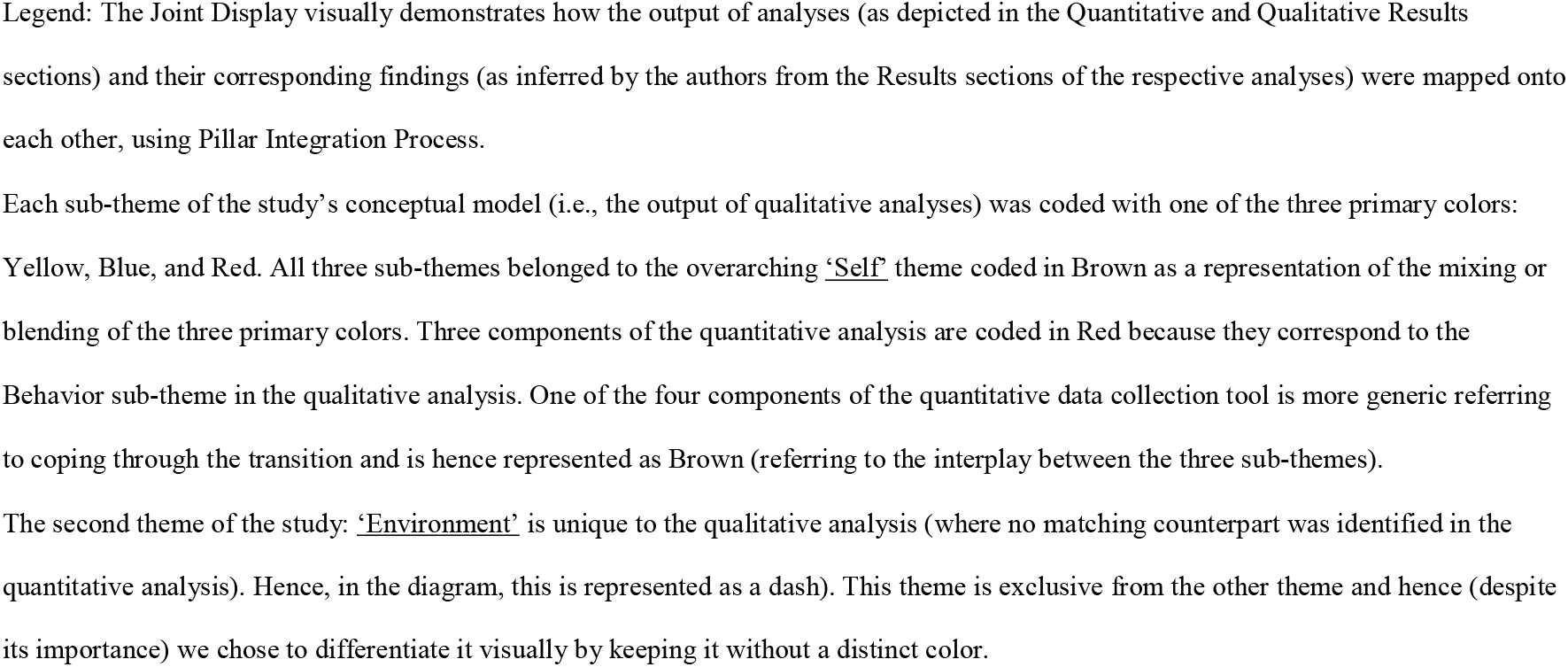
The study’s joint display (merging of the quantitative with the quantitative output of analyses)

## Discussion

This study reinforced the notion that students are self-regulated agents who can manage their learning (31). It also taps into the roles that positive and negative emotions play in SRL (32), and the belief (initially suggested by socio-cognitive theory) that individuals acquire knowledge by observing others and social interactions (33, 34). Both groups of stakeholders perceived themselves to have adapted quite well to change. Also, the instructors perceive themselves to be at an advantage relative to the learners in terms of adapting to the abrupt change induced by COVID-19.

This study demonstrated SRL as a cornerstone in the adaptation to the accelerated change (accompanying COVID-19) in health professions’ education of the individual learners and their instructors. Both groups of postgraduate dental education stakeholders (the learners and instructors) experienced and reported variation in their cognitions, emotions, and behaviors. There was an evident interplay between these individual-level variables, which enabled the stakeholders to adapt to the abrupt change. The stakeholders in the current study also highlighted attributes of the environment that they believe have played a role in their adaptation processes. Along those lines, a recently conducted cross-sectional study aimed at investigating the associations between adaptability, personality, and levels of learning (affective, cognitive, and behavioral) among university students, revealed that adaptability to the pandemic was associated broadly with more positive reactions across multiple indicators (10). This study emphasized the role of adaptability in learning. It appeared that adaptability is acting as a mediator in the association between personality characteristics (i.e., openness, conscientiousness, extraversion, agreeableness, and neuroticism) and students’ reactions to online learning. The more adaptable the students perceived themselves to be, the more constructive were the learning experiences that they reported upon. Moreover, in the same study, the students who reported greater feelings of belonging and mattering perceived themselves to be more adaptable and reported more positive reactions to the learning experiences. This finding highlights the importance of ‘connection to others’ when it comes to online learning. Hence, it is essential for university-level mental health promotion activities to strive to foster adaptability and build ‘academic resilience’, among students, through tapping into elements of self and identity (i.e., internal resources). It is of utmost importance for the students to feel that they belong and matter to other people (i.e., external resources).

The inclusion of instructors in the SRL that takes place in the process of adapting the learning and teaching to accelerated change is not unique to this study. It was previously suggested that the experiential learning that health professions students go through usually entails “strong emotions”. A key responsibility of facilitators of such learning experiences is to harness an environment of trust, authenticity, mutual respect, and integrity (35). This, according to Brookfield (1995), requires educators to be “adult learners”, above all, continuously updating, expanding, and deepening their professional perspectives both on their roles and responsibilities and in relation to the subject matter. He reminds educators that they are required to revisit and analyze their own “visceral” experiences by virtue of their profession before asking their students to do so (36).

The evidence-driven conceptual framework generated from this study confirms the commonality across existent models of SRL and builds upon them (22). These models shed light on the aspects of the self that come together to enable SRL. These include cognitive, metacognitive, behavioral, motivational, and emotional aspects. Similarly, this study highlights the interplay between cognitions (involving metacognitions), emotions (including motivation), and behaviors, along with emphasizing the effect of the all-encapsulating environment. It offers insight into the context: variables that enabled the individuals’ adaptation and those that impeded it. It was previously highlighted that contextual factors impact how students feel a sense of relatedness to their colleagues and instructors (10). All this aligns with the triadic analysis of SRL, which focuses on the relationship between the person, behavior, and environment (37). From the constructivism perspective of experiential learning (i.e., learning through reflection on experience), individuals construct their knowledge through interactions with their environments (35). Individuals in this study appeared to be continuously receiving information from the context and adapting their strategies accordingly. All these insights can be leveraged by other postgraduate dental schools to proactively raise the level of adaptability of individuals, and to create environments that are malleable and conducive to learning goals.

This study shows that the tailor-made quantitative tool is internally reliable and externally valid in the context of this study. There are several tools published in the literature that are designed to measure self-reported adaptation and/ or coping with change including the Coping Strategy Questionnaire (CSQ)(38), Coping Orientation to Problems Experienced (COPE) inventory (39), and its abbreviated version, the Brief COPE (40).

These tools proved of great usefulness across several contexts yet are considerably thorough and time-consuming. There are relevant tools that are more succinct in measuring resilience such as the Brief Resilient Coping Scale (BRCS) (41) and the Brief Resilience Scale (BRS) (42). Yet, none are contextualized to accelerated changes to medical education (including but not limited to the abrupt transition to the online environment) during critical times (such as COVID-19), and factor qualitative reflections into the equation. Accordingly, this study bridges this gap by introducing a concise data collection tool that directly taped into how postgraduate dental students regulated themselves and maneuvered through the exceptionally VUCA environment of COVID-19. Also, the instrument developed and deployed in the current study inquired for qualitative data, which encouraged participants to reflect on their experience. This proved to be of great added value to better understand the processes that the stakeholders went through to self- and co-regulate and in turn thrive.

It is worth noting in terms of the participants’ self-awareness, the modifications in behaviors were the most obvious and easiest to identify in this study. Along those lines, in another exploratory study during COVID-19 (6), modifications in learning (among the learners) or teaching (among the instructors) were also apparent, where learners and instructors modified their approaches to adapt to the rapid transition to distance learning. Such findings that offer insight into the organic growth and development that is inherent to the adaptation process constitute empirical evidence supporting Zimmerman’s cyclical model, which suggests that SRL is a process that involves forethought, followed by performance, and finally, self-reflection (43). Moreover, as evident in the qualitative exploration integral to this study, negative thoughts and emotions surfaced for the stakeholders, whether they were aware of it or not. It is worth highlighting, over here, the importance of building an environment where individuals feel safe to experience, and in turn, let go of these emotions (44). The fact that such negativity surfaces is not unexpected given the VUCA of the situation. The more we empower higher education stakeholders to acknowledge (i.e., become aware of), accept (and respect), and experience (and in turn process) their emotions and thoughts, the less resistant and more adaptable to change they will become (45, 46). This is directly related to fostering cognitive flexibility, which is defined as the ability to adapt the cognitive processing strategies to face new, unexpected, and uncertain conditions in the environment (47, 48)

The current study is characterized by a few caveats that are worth shedding light on. It relied mainly on self-reported data. Each of the two groups of stakeholders provided some reflective data on the other party’s adaptability. It would be interesting for follow-up studies to further explore this point-of-view by systematically enabling observer rating. This will allow for comparing how the perception of one’s adaptability maps onto how others perceive one’s adaptability. Moreover, in alignment with the principles of the Institutional Research function at MBRU, complete anonymity of the participating university stakeholders was maintained. Therefore, demographic details of the participants (e.g., gender and age) or that relating to their affiliation with the university (e.g., year of study and academic title) were not recorded. It would have been interesting to investigate the association between the stakeholders’ extent of adaptability and those variables. Also, although this study offered a lot of insight into how the stakeholders perceive themselves and each other when it comes to adaptability, its cross-sectional design did not enable investigating causality. Hence, it will be great for future studies to be based on longitudinal designs, where potential antecedents to adaptability are captured. The findings of such studies will have substantial practical implications where higher education decision-makers will get a better grasp as to what variables they can foster to proactively raise the level of adaptability among their stakeholders. The study offered a lot of value through the open-ended questions integral to the survey in terms of exploration. Yet, we believe it is worth deploying alternative data collection tools (e.g., focus group sessions) to develop a more thorough understanding of the adaptability experiences of the stakeholders.

## Conclusion

This study encourages opinion leaders in higher education institutions to leverage SRL theories to proactively inspire and empower the learners and their instructors. It also reveals the importance of developing and maintaining safe and nurturing learning environments that foster connection and mattering (to one another), enable cognitive flexibility, and build academic resilience.

